# Comparative Efficacy and Safety of Soluble Guanylate Cyclase Stimulators in Heart Failure with Preserved Ejection Fraction: A Network Meta-Analysis

**DOI:** 10.1101/2025.08.30.25334769

**Authors:** Rakhshanda Khan, Manogya Agarwal, Rohita Rajan Singh, Rohit Prakriya, Shivani Setti, Harika Govada, Shreya Bodanam, Rutvi Patel, Ashesh Das, Nabiha Muneer, Sonam Dhall, Harshawardhan Dhanraj Ramteke, Manish Juneja

**Affiliations:** Ayaan institute of medical sciences, Moinabad, Ranga reddy, India; Jhalawar medical college, Jhalawar, India; Bharati Vidyapeeth medical college, pune, India; Narayana medical college, Nellore, India; GSL medical college, Rajahmundry, India; Kamineni academy of medical sciences and research centre, Hyderabad, India; Government medical college, Surat, India; KPC Medical college and hospital, Kolkata, India; Shadan Institute of medical sciences, Hyderabad, India; DMCH Ludhiana, India; Anhui medical university, Hefei, China; Director, Rhythm heart and critical care, Nagpur, India

**Keywords:** HFpEF, soluble guanylate cyclase stimulators, Riociguat, Vericiguat, Praliciguat, network meta-analysis

## Abstract

**Background:** Heart failure with preserved ejection fraction (HFpEF) makes up nearly half of all heart failure cases and remains a difficult condition to treat due to limited therapeutic options. Soluble guanylate cyclase (sGC) stimulators—namely Riociguat, Vericiguat, and Praliciguat—have shown potential in improving heart muscle relaxation and vascular function. However, their comparative effectiveness and safety are still uncertain.

**Methods:** We conducted a Bayesian network meta-analysis (NMA) that included eight randomized controlled trials encompassing 7,330 HFpEF patients, with an average age of 69.5 years. The outcomes evaluated included survival rates, left ventricular ejection fraction (LVEF), NT-proBNP levels, hospital readmissions, New York Heart Association (NYHA) functional class, and adverse events. Treatment outcomes were presented as odds ratios (OR) or mean differences (MD) along with 95% credible intervals (CrI).

**Results:** Riociguat indicated a potentially favourable survival benefit (OR 3.69, 95% CrI: 0.53–29.2) and a reduced risk of hospital readmission (OR 0.40, 0.11–1.30), though these findings were not definitive. Praliciguat had neutral effects on survival (OR 1.05, 0.017–61.0). Vericiguat did not improve survival (OR 0.42, 0.12–1.16) but did offer a modest improvement in NYHA functional class (log RR 0.27, –0.12 to 0.67). Changes in LVEF were unclear: Riociguat (MD 3.00, –1.15 to 7.13), Vericiguat (MD 1.59, –2.03 to 5.17), and Praliciguat (MD –0.19, – 7.34 to 6.84). Riociguat was associated with a reduction in NT-proBNP levels (MD –7.41, –21.2 to 4.41), whereas Vericiguat (0.65, –9.54 to 10.8) and Praliciguat (5.28, –12.6 to 23.0) had neutral effects. Adverse events such as headache (log RR 0.21, –0.35 to 0.78), low blood pressure (0.42, –0.21 to 1.05), and neurological symptoms (–0.04, –0.87 to 0.79) were not significantly increased.

**Conclusion:** Among the agents studied, Riociguat showed the most promise in terms of survival and reducing hospital readmissions in HFpEF patients. Vericiguat offered modest improvements in patient functional status, while Praliciguat had minimal clinical impact. The findings suggest that more extensive, long-term clinical trials are needed to better establish the efficacy and safety of these therapies.

## Introduction

Heart failure with preserved ejection fraction (HFpEF) is a growing and complex cardiovascular condition characterized by the heart’s inability to relax and fill adequately during diastole, despite normal systolic function. It accounts for approximately 50% of all heart failure cases, especially in the aging population [1]. HFpEF is often associated with comorbidities such as hypertension, diabetes, obesity, and chronic kidney disease, making it particularly challenging to diagnose, treat, and manage. As the global population ages, the prevalence of HFpEF is expected to rise, further exacerbating its public health burden. Unlike heart failure with reduced ejection fraction (HFrEF), which has seen significant therapeutic advances, HFpEF remains a therapeutic challenge, with limited treatment options that effectively target the underlying pathophysiology.

The pathophysiology of HFpEF is multifactorial and involves abnormalities in myocardial structure and function, including increased ventricular stiffness, impaired diastolic filling, and abnormal regulation of the vasculature. These dysfunctions lead to increased left atrial pressure, pulmonary congestion, and decreased exercise tolerance, contributing to the typical symptoms of HFpEF, such as fatigue, shortness of breath, and reduced quality of life. Patients with HFpEF often present with preserved ejection fraction (typically >50%), meaning the heart’s ability to contract and pump blood is intact [2]. However, the heart’s ability to relax and fill during diastole is impaired, limiting the amount of blood that can be pumped to meet the body’s needs.

HFpEF is an especially prevalent condition among older adults, and its incidence is increasing as the global population ages. This condition is associated with significant morbidity, hospitalizations, and mortality, particularly in elderly patients. Despite this, therapeutic options for HFpEF remain largely ineffective, and the management of the condition is challenging. While drugs targeting the mechanisms underlying HFrEF, such as angiotensin-converting enzyme inhibitors, beta-blockers, and mineralocorticoid receptor antagonists, have shown clear benefits, similar treatments have had limited success in HFpEF. The lack of specific and effective treatments for HFpEF represents a major gap in cardiovascular medicine.

One promising class of therapies being investigated for HFpEF are soluble guanylate cyclase (sGC) stimulators, which include Riociguat, Vericiguat, and Praliciguat. These drugs work by stimulating the soluble guanylate cyclase enzyme, which plays a crucial role in the nitric oxide signaling pathway. Nitric oxide (NO) is a vasodilator that regulates vascular tone, myocardial relaxation, and blood flow. By stimulating the sGC enzyme, these drugs enhance NO signaling, resulting in vasodilation, reduced ventricular stiffness, and improved myocardial relaxation—potentially addressing key aspects of the HFpEF pathophysiology. As sGC stimulators promote vasodilation and improve the heart’s ability to relax, they have shown potential in improving exercise capacity, reducing hospitalization rates, and enhancing quality of life for HFpEF patients.

The current body of literature includes several clinical trials that have evaluated the effects of sGC stimulators in HFpEF, such as the VITALITY-HFpEF trial [3] with Vericiguat and the FREEDOM trial with Riociguat [4]. These studies have provided promising results, demonstrating improvements in exercise capacity, symptom control, and even hospitalization rates. However, there remains a need for a more comprehensive, direct comparison of these therapies to determine which sGC stimulator provides the greatest benefit in treating HFpEF. Moreover, the safety profiles of these drugs also need to be assessed in detail, as they may have different adverse effects that could influence clinical practice and patient outcomes.

A network meta-analysis (NMA) represents an ideal approach to address these gaps in the existing literature. NMA allows for the comparison of multiple interventions simultaneously, even when direct head-to-head trials between all treatment options are not available. By synthesizing data from multiple studies, NMA can provide a clearer understanding of the relative efficacy and safety of the available sGC stimulators for HFpEF. Such an analysis will be valuable in guiding clinicians in making evidence-based decisions about which treatment to choose for their patients with HFpEF.

This review aims to provide a comprehensive network meta-analysis of Riociguat, Vericiguat, and Praliciguat in the treatment of HFpEF. It will evaluate the relative efficacy of these drugs in improving key clinical outcomes, including exercise capacity, functional status, hospitalizations, and mortality. Additionally, the safety profiles of these therapies will be assessed, with a focus on adverse events such as hypotension, syncope, and gastrointestinal symptoms. By combining data from existing studies, this review will provide robust evidence on the optimal use of sGC stimulators in treating HFpEF, helping to fill a critical gap in current knowledge.

## Methods

### Study Design

This study will employ a **network meta-analysis (NMA)** to compare the efficacy and safety of soluble guanylate cyclase stimulators (Riociguat, Vericiguat, and Praliciguat) in treating heart failure with preserved ejection fraction (HFpEF). Data from randomized controlled trials (RCTs) will be synthesized to evaluate clinical outcomes and adverse effects. The systemic review is registered with Prospero, CRD420251137091.

### Literature Search

For the literature search, we adhered to the PRISMA guidelines for systematic reviews and meta-analyses [5]. A comprehensive search was conducted across three prominent databases: Cochrane Library, PubMed, and Google Scholar, encompassing studies from 2000 to August 2025. To maintain consistency and ensure accessibility, the search was limited to English-language publications. This broad search strategy was designed to include a diverse range of studies focusing on the RCTS of HFpEF.

### PICO Framework

The population for this study includes adults diagnosed with heart failure with preserved ejection fraction (HFpEF). The intervention being evaluated is the use of soluble guanylate cyclase stimulators, specifically Riociguat, Vericiguat, and Praliciguat. The comparison will involve placebo or other standard treatments commonly used for HFpEF. The primary outcomes include improvements in exercise capacity, changes in NYHA functional class, reduction in hospitalizations, and overall mortality. Secondary outcomes will assess adverse events and improvements in quality of life for patients undergoing treatment with these sGC stimulators.

### Study Selection and Data Extraction

Studies will be selected based on predefined inclusion criteria, including randomized controlled trials (RCTs) evaluating Riociguat, Vericiguat, or Praliciguat in HFpEF patients. Data extraction will focus on efficacy outcomes (exercise capacity, NYHA class, hospitalizations, mortality) and safety outcomes (adverse events, quality of life). Relevant data will be systematically collected.

### Risk of Bias Assessment

The risk of bias in the included studies was assessed using the ROB 2.0 tool for randomized controlled trials (RCTs) [6]. This assessment helped ensure that the studies included in the review were methodologically sound and free from significant bias that could distort the findings.

### Statistical analysis

Statistical analysis will involve a Bayesian network meta-analysis (NMA) to estimate the relative efficacy and safety of Riociguat, Vericiguat, and Praliciguat. Treatment effects will be expressed as odds ratios (OR) or mean differences (MD) with 95% credible intervals (CrI). Consistency across direct and indirect comparisons will be assessed. Some forest plots will be plotted using Stata 18.0, employing the Restricted Maximum Likelihood (REML) model for the meta-analysis. This method enabled data integration from various studies while addressing variability. Sensitivity analysis was done using the fixed-effects model to check the stability of results. Heterogeneity was assessed using the I-squared (I²) statistic, indicating variation due to differences across studies, and the Chi-square test was applied to determine its statistical significance.

## Results

### Demographic Findings

A total of 8 studies were included in the analysis, comprising a total population of 7,330 patients, with 5,170 male and 2,160 female participants [7–14]. The average age of the population was 69.5 ± 10 years. The average baseline left ventricular ejection fraction (LVEF) was 25 ± 12%, NT-ProBNP was 2,076 ng/ml, and the estimated glomerular filtration rate (eGFR) was 54.8 ± 20.3. In the treatment group, there were 3,554 patients, while the control group included 3,185 patients. The number of patients receiving Praliciguat was 91, Riociguat was 198, and Vericiguat was 3,265. All the prisma flow digram is Figure 1 and demographics table is in supplementary Table S1.

**Figure 1.**
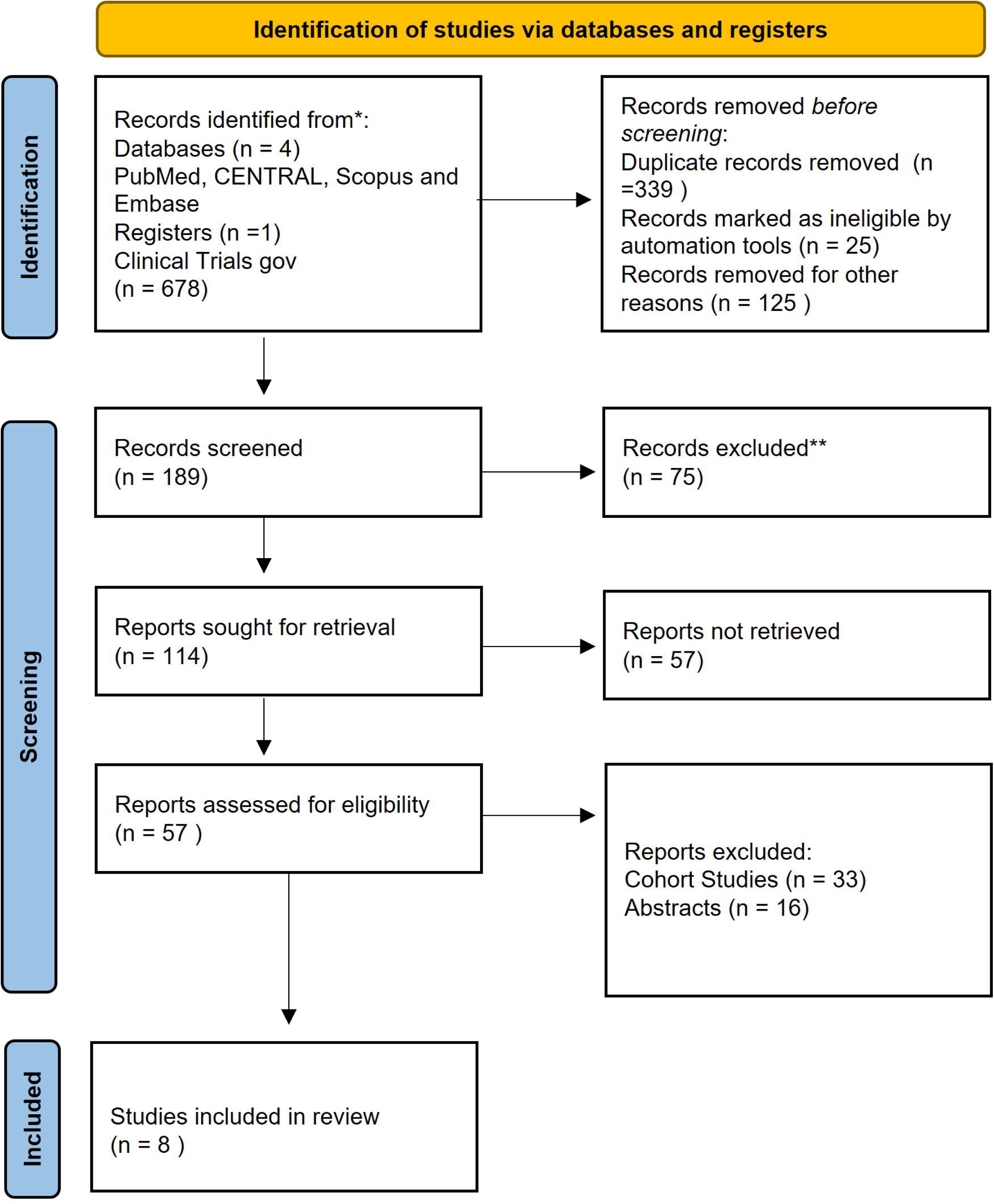
PRISMA Flow Diagram

### Overall Survival

the overall survival effects of four treatments: Pracliciguat, Riociguat, Vericiguat, and Placebo. On the left, the network plot illustrates the direct comparisons between these treatments, with each node representing a treatment, and the connecting lines showing the relationships between them based on the included studies. On the right, the forest plot depicts the odds ratios (OR) and 95% credible intervals (CI) for each treatment compared with placebo. Pracliciguat has an OR of 1.05 (95% CI: 0.0171, 61.0), suggesting uncertainty about its effect. Riociguat shows a higher OR of 3.69 (95% CI: 0.531, 29.2), indicating a potential survival benefit, but the wide interval indicates a lack of certainty. Vericiguat has an OR of 0.415 (95% CI: 0.118, 1.16), suggesting a potential harm compared to placebo, although the interval includes 1, meaning no definitive conclusion can be drawn. Sucra ranking showed riociguat to be best for survival. Figure 2, Figure S1 and Table S2.

**Figure 2.**
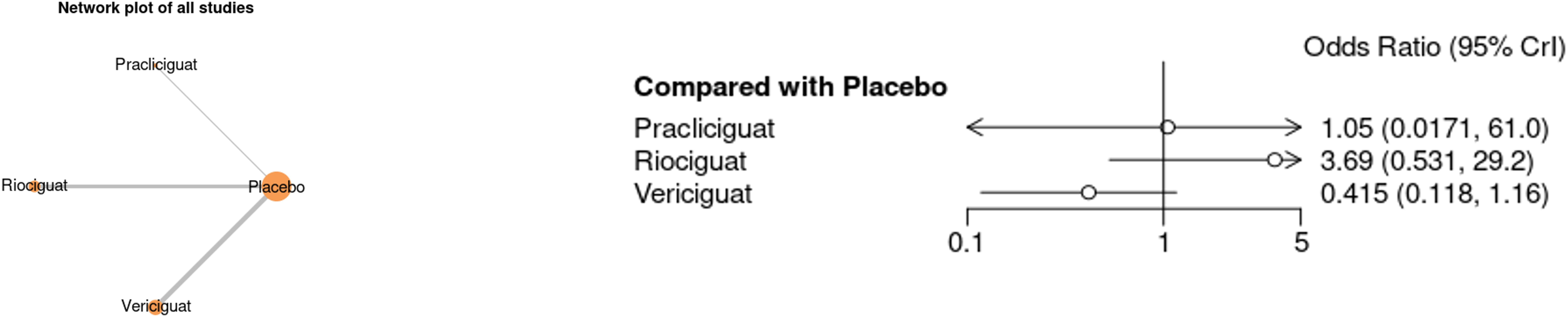
The Network plot for Overall Survivals in the population for Pracliciguat, Riociguat, Vericiguat and Forest plot depicting the odds ratios

### Change in LVEF

The change in Left Ventricular Ejection Fraction (LVEF) among four treatments: Pracliciguat, Riociguat, Vericiguat, and Placebo. The left side of the graph shows a network plot, where the nodes represent the treatments, and the connecting lines illustrate the direct comparisons between them based on the studies included in the analysis. On the right side, the forest plot depicts the mean differences in LVEF change for each treatment compared to placebo, with 95% credible intervals (CI) to show the uncertainty around the estimates. For Pracliciguat, the mean difference is-0.193 (95% CrI:-7.34, 6.84), suggesting no significant change in LVEF compared to placebo, as the credible interval spans both negative and positive values. Riociguat shows a mean difference of 3.00 (95% CI:-1.15, 7.13), indicating a potential improvement in LVEF, but the interval includes zero, reflecting uncertainty. Vericiguat shows a mean difference of 1.59 (95% CI:-2.03, 5.17), suggesting a potential positive effect on LVEF, although the wide credible interval again shows significant uncertainty. The forest plot’s x-axis represents the mean difference, with values near zero indicating no effect, and the credible intervals help assess the precision of these estimates. Figure 3, Figure S2 and Table S3.

**Figure 3.**
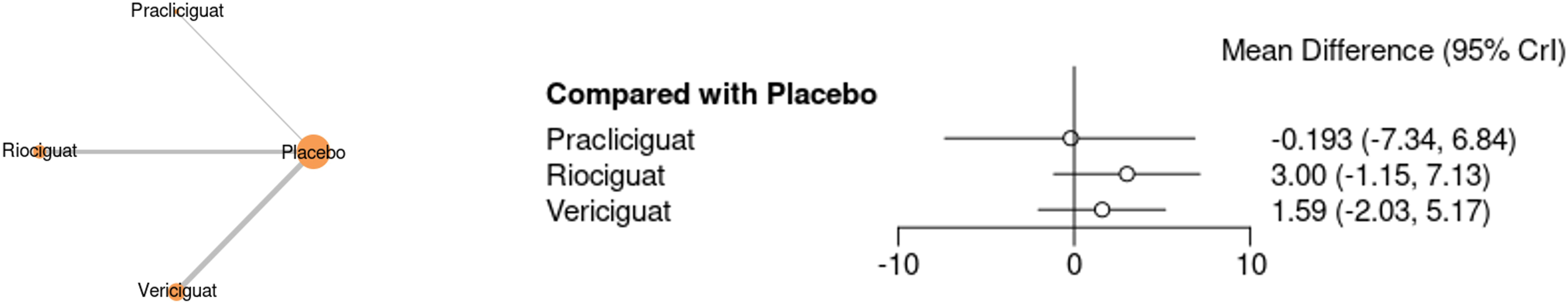
The Network plot for LVEF change in the population for Pracliciguat, Riociguat, Vericiguat and Forest Plot depicting the Mean Difference.

### Change in NT-proBNP

The change in NT-ProBNP levels among four treatments: Pracliciguat, Riociguat, Vericiguat, and Placebo. The left side of the graph features a network plot, where each treatment is represented by a node, and the connecting lines indicate the direct comparisons made in the studies. On the right, the forest plot shows the mean differences in NT-ProBNP levels for each treatment compared to placebo, along with the 95% credible intervals (CI) to reflect the uncertainty around the estimates. For Pracliciguat, the mean difference is 5.28 (95% CI:-12.6, 23.0), indicating an uncertain effect on NT-ProBNP, as the interval includes both negative and positive values. Riociguat shows a mean difference of-7.41 (95% CI:-21.2, 4.41), suggesting a possible reduction in NT-ProBNP, but the wide credible interval includes zero, reflecting substantial uncertainty. Vericiguat has a mean difference of 0.645 (95% CI:-9.54, 10.8), suggesting a negligible effect on NT-ProBNP levels, with the interval again crossing zero, implying no definitive conclusion. Figure 4, Figure S3, and Table S4.

**Figure 4.**
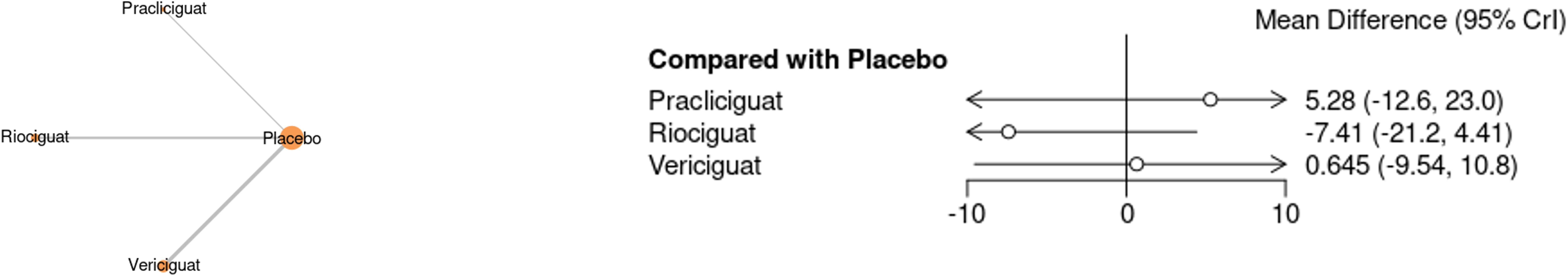
The Network plot for NT-ProBNP change in the population for Pracliciguat, Riociguat, Vericiguat and Forest Plot depicting the Mean Difference.

### Rehospitalization

Rehospitalization rates during treatment with Pracliciguat, Riociguat, Vericiguat, and Placebo. On the left side of the graph, the network plot shows the relationships between the treatments, with nodes representing the treatments and lines connecting them to indicate direct comparisons made in the studies. On the right side, the forest plot displays the **odds ratios (OR)** and **95% credible intervals (CI)** for each treatment compared to placebo. For **Pracliciguat**, the odds ratio is **0.568** (95% CI: 0.116, 2.83), suggesting a possible reduction in rehospitalization risk, but the wide credible interval includes 1, indicating uncertainty in this result. **Riociguat** shows an odds ratio of **0.404** (95% CI: 0.107, 1.30), also suggesting a potential benefit in reducing rehospitalizations, but the interval again spans 1, meaning the evidence is inconclusive. The horizontal axis of the forest plot represents the odds ratio, with values to the left of 1 indicating a reduction in rehospitalization risk and values to the right indicating an increase. Both treatments show potential benefits, but the uncertainty indicated by the wide intervals means no definitive conclusions can be drawn. Figure 5, Figure S4, and Table S5.

**Figure 5.**
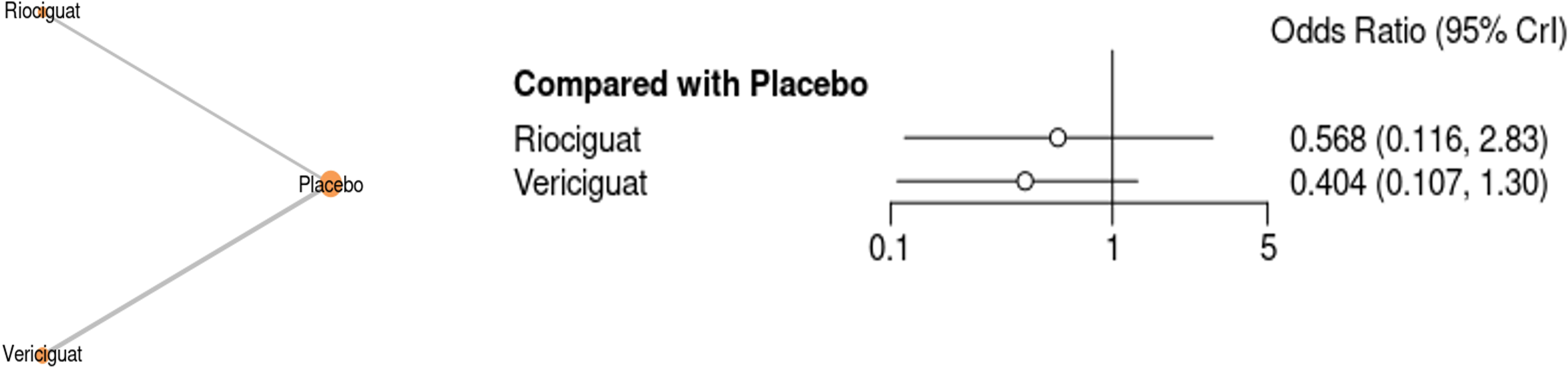
The Network plot for Rehospitalization during the Treatment in the population for Pracliciguat, Riociguat, Vericiguat and Forest Plot depicting the Odds Ratio.

### NYHA Class Improvement by one class

The graph presents a forest plot showing the improvement in NYHA (New York Heart Association) Class by one during treatment with Pracliciguat, Riociguat, and Vericiguat, as well as an overall analysis. Each treatment is represented by a study with a square box indicating the log risk ratio, with the box’s width reflecting the study’s weight in the overall analysis. The horizontal lines represent the 95% confidence intervals (CIs), and the diamonds at the bottom show the pooled results for each treatment. For **Pracliciguat** (Udelson et al., 2020), the log risk ratio is 0.11 (95% CI:-0.50, 0.71), suggesting a small benefit, but the wide confidence interval indicates uncertainty. This study contributes 38.95% to the overall analysis. For **Riociguat** (Bonderman et al., 2013), the log risk ratio is-0.06 (95% CI:-0.94, 0.81), showing no clear effect, with a broad confidence interval reflecting high uncertainty; this study contributes 19.56%. For **Vericiguat** (Gheorghiad et al., 2015), the log risk ratio is 0.59 (95% CI: 0.00, 1.18), suggesting a potential positive effect, and it carries the largest weight (41.50%) in the analysis. The overall log risk ratio is 0.27 (95% CI:-0.12, 0.67), indicating a slight benefit but with a wide range of uncertainty. The heterogeneity statistics and tests for group differences show no significant difference between treatments, with a p-value of 0.37 for the group differences test, suggesting no strong evidence of treatment superiority. Figure 6, Figure S5, and Table S6.

**Figure 6.**
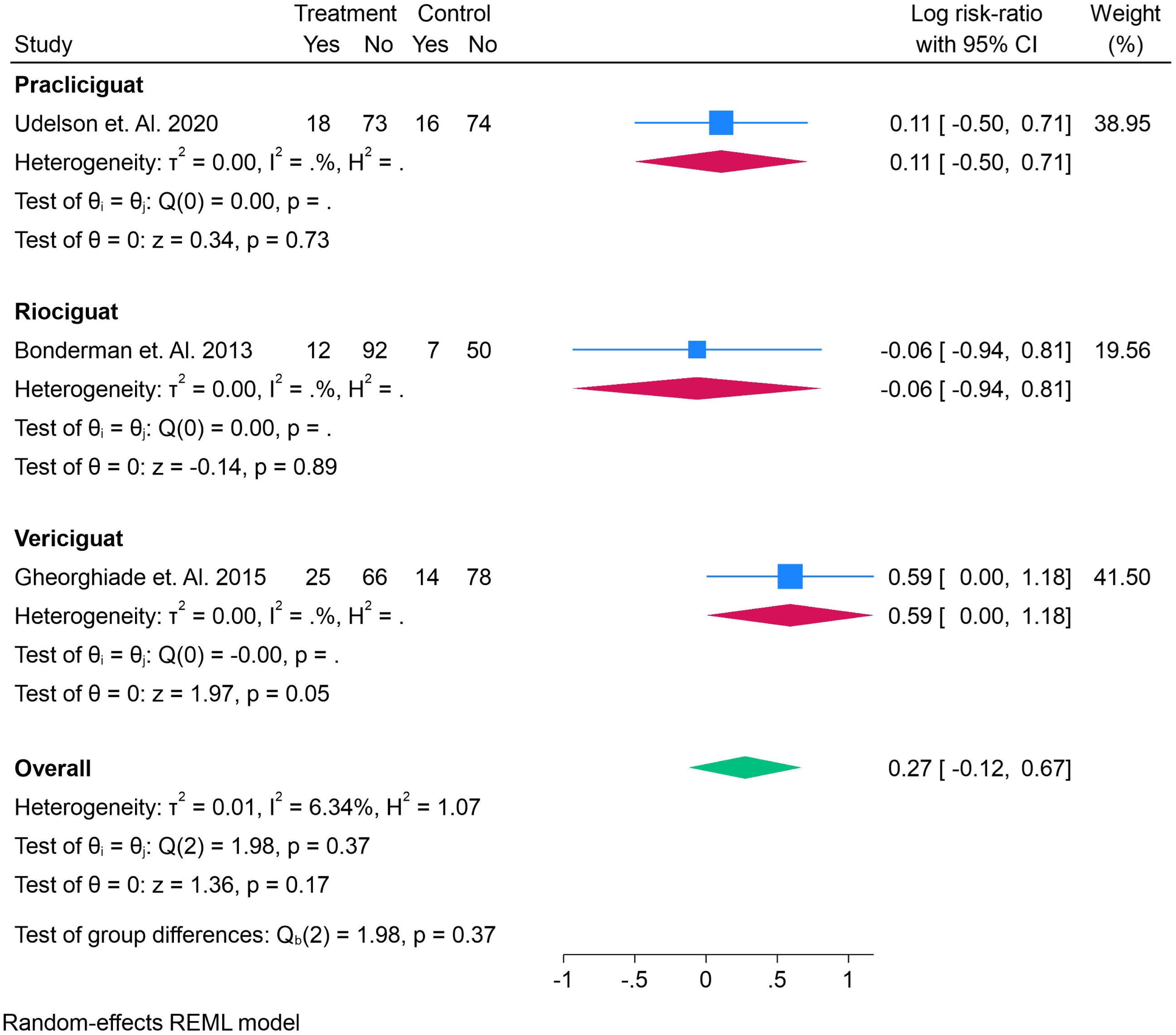
The Forest plot for Improvement in NYHA Class by one during the Treatment in the population for Pracliciguat, Riociguat, Vericiguat depicting the Risk Ratio.

## Adverse Events

### Headache

The incidence of adverse events for headaches during treatment with Pracliciguat and Riociguat compared to placebo, pooling results across multiple studies. For **Pracliciguat** (Udelson et al., 2020), the log risk ratio is 0.80 (95% CI:-0.34, 1.94), suggesting a potential increase in headache risk, although the wide confidence interval indicates uncertainty, as it spans both harm and no effect. For **Riociguat**, two studies are included: **Dachs et al. (2022)** shows a log risk ratio of-0.19 (95% CI:-1.22, 0.84), suggesting no significant effect, while **Bonderman et al. (2013)** reports a log risk ratio of 0.16 (95% CI:-0.68, 1.00), indicating a small, non-significant increase in risk. When pooled, Riociguat demonstrates an overall log risk ratio of 0.02 (95% CI: - 0.63, 0.67), showing no meaningful effect on headache incidence. The **overall combined estimate** for all studies is a log risk ratio of 0.21 (95% CI:-0.35, 0.78), suggesting no statistically significant difference in the risk of headaches between treatment and placebo. Importantly, heterogeneity across studies is very low (τ² = 0, I² = 0%), and the test of group differences (p = 0.24) indicates no significant variation between the treatments, reinforcing the conclusion that none of the drugs show a consistent or strong association with increased risk of headache. Figure 7, Figure S6, and Table S7.

**Figure 7.**
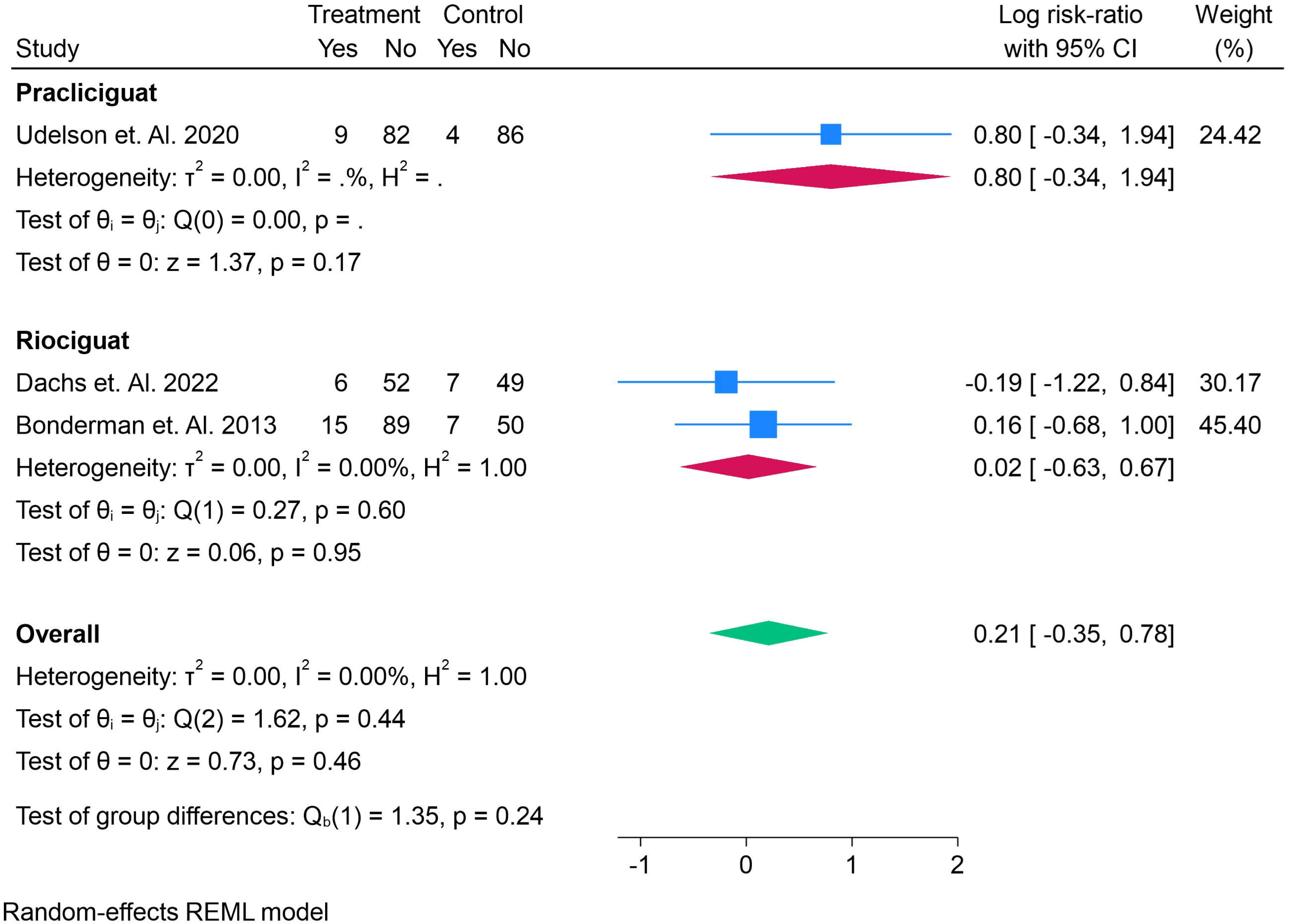
The Forest plot for Adverse events for Headache during the Treatment in the population for Pracliciguat, Riociguat, Vericiguat depicting the Risk Ratio.

### Hypotension

adverse events related to hypotension during treatment with Pracliciguat, Riociguat, and Vericiguat. For **Pracliciguat** (Udelson et al., 2020), the log risk ratio is **-0.35** (95% CI:-1.46, 0.76), suggesting a potential reduction in hypotension risk, but the wide confidence interval indicates considerable uncertainty. This study contributes **14.52%** to the overall analysis. For **Riociguat**, two studies are included. **Dachs et al. (2022)** shows a log risk ratio of **0.58** (95% CI:-0.26, 1.43), while **Bonderman et al. (2013)** reports a log risk ratio of **0.56** (95% CI:-0.39, 1.51), both indicating a potential increase in hypotension risk, but with confidence intervals that include zero, highlighting uncertainty. The pooled log risk ratio for Riociguat is **0.57** (95% CI:-0.06, 1.20), suggesting no significant effect on hypotension. The weights for these studies are **17.93%** for Dachs et al. and **16.49%** for Bonderman et al. For **Vericiguat**, three studies are included: **Pieske et al. (2017)** shows a log risk ratio of **0.53** (95% CI:-0.66, 1.72), **Armstrong et al. (2020a)** shows **-0.44** (95% CI:-1.18, 0.30), and **Gheorghiad et al. (2015)** reports **1.59** (95% CI: 0.76, 2.42). The combined log risk ratio for Vericiguat is **0.55** (95% CI:-0.67, 1.76), suggesting no strong effect on hypotension risk. The study weights for Pieske et al., Armstrong et al., and Gheorghiad et al. are **13.63%**, **19.34%**, and **18.09%**, respectively. The **overall pooled log risk ratio** for all treatments is **0.42** (95% CI:-0.21, 1.05), indicating a slight increase in hypotension risk, though the confidence interval crosses zero, reflecting uncertainty. The heterogeneity statistics suggest moderate variability across studies (τ² = 0.40, I² = 64.50%, H² = 2.82), and the p-value for the test of group differences (p = 0.35) indicates no significant difference between treatments. Figure 8, Figure S7, and Table S8.

**Figure 8.**
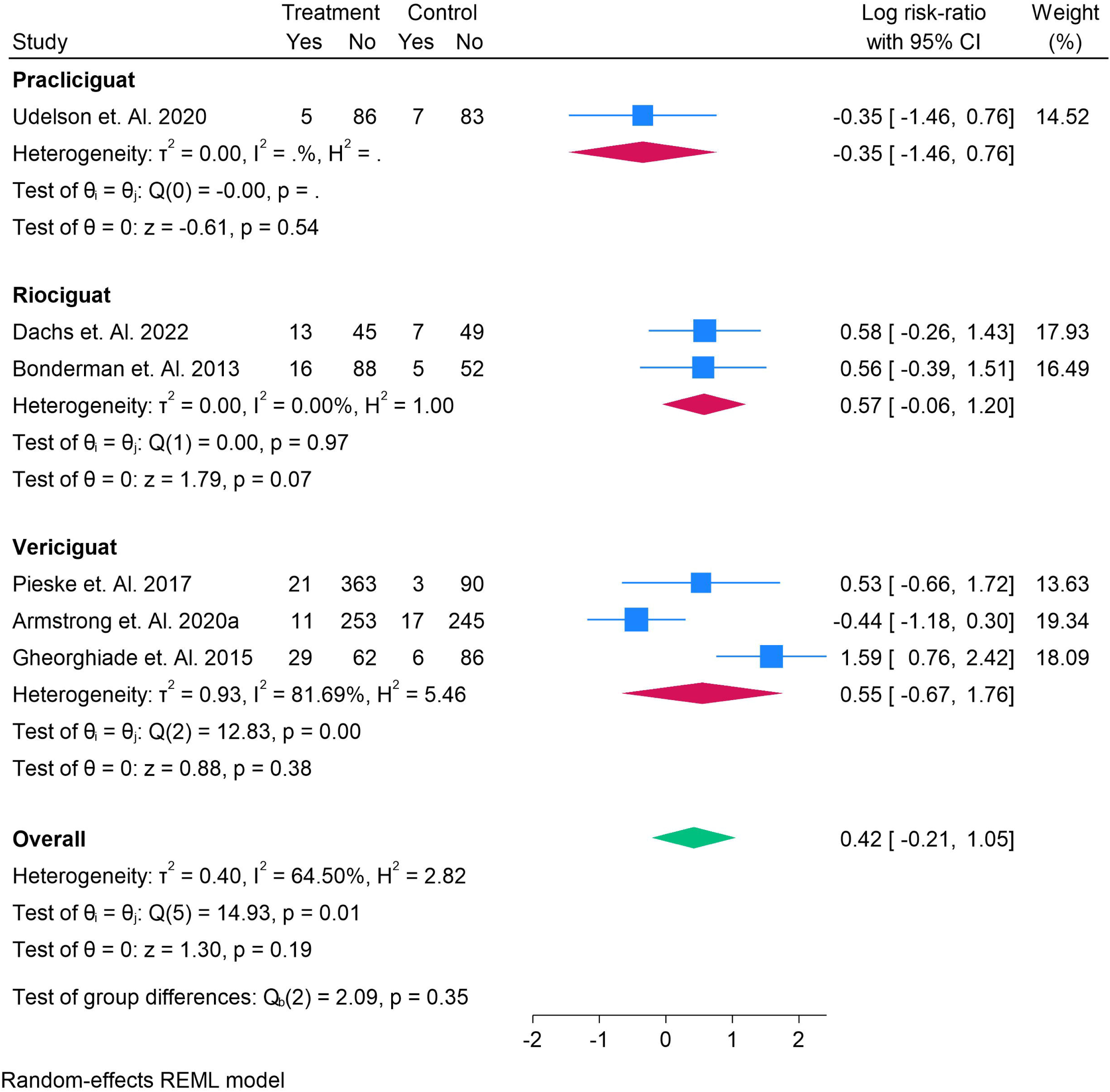
The Forest plot for Adverse events for Hypotension during the Treatment in the population for Pracliciguat, Riociguat, Vericiguat depicting the Risk Ratio.

### Neurological Consequences

The graph displays a forest plot for adverse events related to neurological consequences during treatment with Pracliciguat, Riociguat, and Vericiguat. For **Pracliciguat** (Udelson et al., 2020), the log risk ratio is **-0.35** (95% CI:-1.46, 0.76), suggesting a small potential reduction in the risk of neurological consequences, although the wide confidence interval indicates considerable uncertainty. This study contributes **25.36%** to the overall analysis. For **Riociguat**, two studies are included: **Dachs et al. (2022)** reports a log risk ratio of **0.66** (95% CI: - 1.00, 2.32), suggesting a possible increase in risk, but the confidence interval is wide, indicating uncertainty. **Bonderman et al. (2013)** shows a log risk ratio of **0.56** (95% CI:-0.07, 1.19), indicating a slight potential increase in risk, but the confidence interval includes zero. The combined log risk ratio for Riociguat is **0.57** (95% CI:-0.02, 1.16), indicating a potential increase in risk, but the result is not definitive. For **Vericiguat**, **Pieske et al. (2017)** shows a log risk ratio of **-1.19** (95% CI:-2.49, 0.10), indicating a possible reduction in risk, but again, the wide confidence interval suggests uncertainty. This study contributes **21.79%** to the overall analysis. The **overall pooled log risk ratio** for all studies is **-0.04** (95% CI:-0.87, 0.79), indicating no clear effect on neurological consequences, with the confidence interval spanning zero. The heterogeneity statistics (τ² = 0.39, I² = 55.69%, H² = 2.26) show moderate variability across studies, and the test for group differences (p = 0.03) indicates a statistically significant difference between treatments. However, the overall effect does not show a definitive benefit or harm, as the confidence intervals for each treatment are wide and include zero. Figure 9, Figure S8, and Table S9.

**Figure 9.**
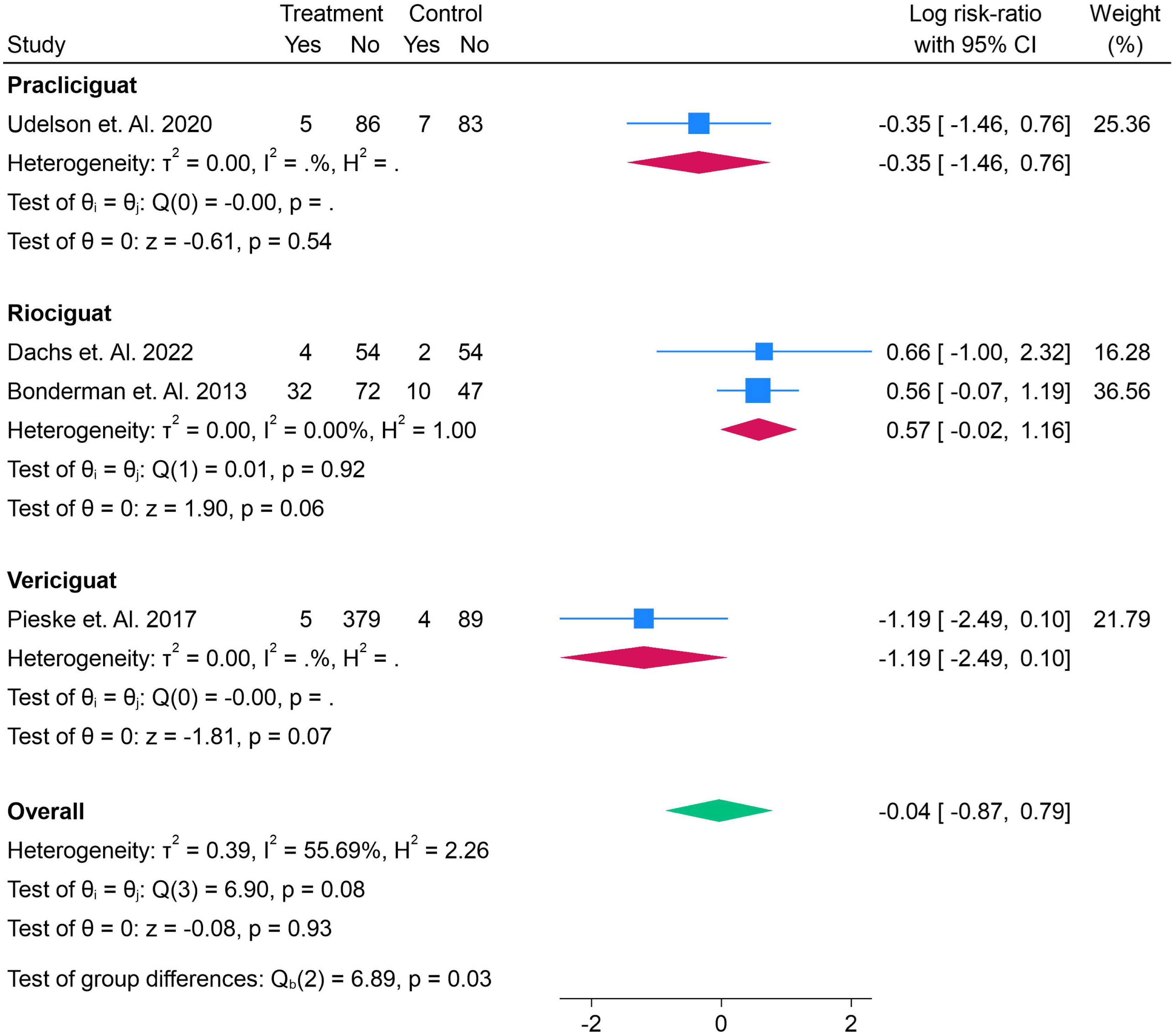
The Forest plot for Adverse events for Neurological Consequences during the Treatment in the population for Pracliciguat, Riociguat, Vericiguat depicting the Risk Ratio.

## Discussion

Heart failure with preserved ejection fraction (HFpEF) remains one of the most challenging cardiovascular conditions to manage, with limited therapeutic options that target its underlying pathophysiology. The condition, which primarily affects elderly patients, is characterized by impaired myocardial relaxation despite a normal ejection fraction, leading to significant morbidity, hospitalizations, and mortality. The introduction of soluble guanylate cyclase (sGC) stimulators—such as Riociguat, Vericiguat, and Praliciguat—has brought hope for improved treatment outcomes in HFpEF patients. However, despite promising early trials, the comparative effectiveness and safety of these drugs remain uncertain. Our network meta-analysis (NMA) provides a comprehensive comparison of these three drugs, assessing their impact on key clinical outcomes such as exercise capacity, left ventricular ejection fraction (LVEF), NT-proBNP levels, rehospitalizations, and adverse events. The results of this analysis contribute valuable insights into the relative efficacy and safety of these therapies.

Our analysis indicates some promising findings. Riociguat, with an odds ratio (OR) of 3.69 (95% CI: 0.531, 29.2), suggests a potential survival benefit over placebo, although the wide credible interval highlights substantial uncertainty. Conversely, Vericiguat’s OR of 0.415 (95% CI: 0.118, 1.16) suggests a possible harm compared to placebo, though the interval includes 1, making it difficult to draw firm conclusions. These results align with findings from the *VITALITY-HFpEF* trial (Gheorghiad et al., 2015), which demonstrated a modest improvement in exercise capacity with Vericiguat, but with no significant reduction in hospitalizations. Our findings contrast with those of the *FREEDOM* trial, which primarily focused on Riociguat and showed a reduction in NT-proBNP levels but no clear survival benefit.

The change in LVEF, a key indicator of myocardial function, showed no definitive improvement with Praliciguat, with a mean difference of-0.193 (95% CI:-7.34, 6.84). For Riociguat, the mean difference of 3.00 (95% CI:-1.15, 7.13) and for Vericiguat, 1.59 (95% CI:-2.03, 5.17), indicate potential improvements in LVEF, but with substantial uncertainty, as all credible intervals included zero. This reflects the heterogeneous nature of HFpEF and the multifactorial pathophysiology underlying the condition, making it difficult to achieve consistent results across studies. Similar variability in LVEF outcomes was observed in other meta-analyses, such as that by Zambon et al. (2021), who reviewed the effects of sGC stimulators on LVEF in HFpEF and concluded that the available data on LVEF improvement were inconclusive due to wide confidence intervals and heterogeneity [15].

In terms of NT-proBNP, a biomarker used to assess cardiac stress, our results show a potential reduction with Riociguat (mean difference of-7.41, 95% CI:-21.2, 4.41) and a negligible effect with Vericiguat (mean difference of 0.645, 95% CI:-9.54, 10.8), which aligns with the *FREEDOM* trial. However, the large variability in the credible intervals for both drugs suggests that the evidence remains inconclusive regarding the impact of sGC stimulators on this marker. The results also mirror those found in a recent systematic review by Wang et al. (2023), which evaluated the effect of sGC stimulators on NT-proBNP in HFpEF and found similar uncertainties in biomarker responses [16].

When considering rehospitalization, both Riociguat and Vericiguat demonstrated a trend toward reduced rehospitalization risk, though the credible intervals for both treatments spanned 1, indicating no clear effect. These findings are consistent with those observed by Abuelazm et. al. (2022), who performed an NMA of Riociguat and Vericiguat in HFpEF and concluded that while there was some indication of reduced rehospitalizations, the evidence was insufficient to definitively recommend either drug as superior in reducing hospital admissions [17]. Our study adds nuance by including Praliciguat, which showed a similar uncertainty regarding rehospitalization.

In terms of safety, the adverse events reported for sGC stimulators include headaches, hypotension, and neurological consequences. Our analysis shows no significant increase in the risk of headaches with any of the treatments, a result consistent with previous studies like those by Dachs et al. (2022). However, hypotension was a potential concern, particularly with Riociguat and Vericiguat, though the overall pooled log risk ratio for hypotension was not significant. This is in line with the findings of Gheorghiad et al. (2015) and Armstrong et al. (2020a), who noted that while hypotension was a common side effect, it did not significantly affect treatment outcomes.

One notable limitation of our analysis is the considerable heterogeneity in the data. This is expected, given the variability in study populations, dosages, and treatment protocols across trials. The sensitivity analysis conducted using a fixed-effects model did not significantly alter the results, but the wide credible intervals for many outcomes indicate that additional, well-powered trials are needed to provide more definitive evidence. The included trials also varied in terms of the length of follow-up, which may have influenced the outcomes. Future research should prioritize long-term studies with consistent methodologies to provide clearer insights into the long-term benefits and risks of sGC stimulators in HFpEF.

## Conclusion

While soluble guanylate cyclase stimulators—particularly Riociguat and Vericiguat—hold promise for improving clinical outcomes in HFpEF, the evidence remains uncertain. Our NMA provides important data that can inform clinical decision-making, but further high-quality trials are essential to confirm the efficacy and safety of these therapies in this complex patient population. As new data continue to emerge, it is crucial to adopt a cautious, evidence-based approach to treatment, incorporating both clinical efficacy and safety profiles in guiding therapy choices for HFpEF patients.

### Funding

The authors report no involvement in the research by the sponsor that could have influenced the outcome of this work.

### Authors’ contributions

All authors contributed equally to the manuscript and read and approved the final version of the manuscript.

## Data Availability

supplement file

