## supplementary file for "Comparative Efficacy and Safety of Soluble Guanylate Cyclase Stimulators in Heart Failure with Preserved Ejection Fraction: A Network Meta-Analysis"

Search String

("heart failure" OR "cardiac failure" OR HFrEF OR HFpEF) AND ("guanylate cyclase stimulator" OR riociguat OR vericiguat OR praliciguat OR "sGC stimulator")

Table S1. Demographics and Baseline Characteristics

| **Reference Number** | **Author Year** | **Country** | **Total Population** | **Total Males** | **Total Females** | **Average Age** | **Follow up Duration** | **LVEF** | **NT** | **eGFR** | **HF type** | **Grade** | **Main Finding** |
| --- | --- | --- | --- | --- | --- | --- | --- | --- | --- | --- | --- | --- | --- |
| 7 | Dachs et. Al. 2022 | Austria and Germany | 114 | 31 | 83 | 71.4 | 6 | 60.6 ± 6.3 | 794.1 | 62.6 ± 20.9 | HFpEF | High | Riociguat improved cardiac output in patients with pulmonary hypertension associated with heart failure with preserved ejection fraction (PH-HFpEF), though it did not significantly improve clinical outcomes or quality of life. |
| 8 | Pieske et. Al. 2017 | Global | 477 | 250 | 227 | 73 | 3 | 57 ± 5 | 1174 | 54.8 ± 20.3 | HFpEF | High | Vericiguat was well tolerated but did not significantly affect NT-proBNP or left atrial volume compared to placebo at 12 weeks in patients with HFpEF |
| 9 | Armstrong et. Al. 2020a | Global | 789 | 385 | 404 | 72.7 | 6 | 56 ± 7.9 | 1403 | 59.6 ± 21.2 | HFpEF | High | Vericiguat did not improve the physical limitation score of the KCCQ in patients with HFpEF after 24 weeks compared with placebo. |
| 10 | Udelson et. Al. 2020 | USA and Canada | 196 | 106 | 90 | 70.4 | 3 | 59 ± 7 | 2604 | N/A | HFpEF | High | Praliciguat did not significantly improve peak VO2 in patients with HFpEF |
| 11 | Armstrong et. Al. 2020b | Global | 5050 | 3842 | 1208 | 67.3 | 10.8 | 29 ± 8.3 | 2816 | N/A | HFpEF | High | Vericiguat significantly reduced the incidence of death from cardiovascular causes or hospitalization for heart failure compared to placebo. |
| 12 | Bonderman et. Al. 2014 | Austria and Germany | 47 | 18 | 29 | 75.1 | 6 | 62.5 ± 6.9 | 1747 | N/A | HFpEF | High | Riociguat 2 mg significantly increased stroke volume and cardiac index while reducing systolic blood pressure and right ventricular end-diastolic area, with no significant effect on pulmonary artery pressures |
| 13 | Bonderman et. Al. 2013 | Global | 201 | 172 | 29 | 58.1 | 4 | 27.1 ± 0.6 | 3000 | 68.7 ± 0.6 | HFpEF | High | Riociguat significantly improved cardiac index, pulmonary vascular resistance, and systemic vascular resistance without major changes in pulmonary artery pressure. |
| 14 | Gheorghiade et. Al. 2015 | Europe, North America and Asia | 456 | 366 | 90 | 68 | 3 | 29.6 ± 8.4 | 3076 | 58 ± 19.5 | HFpEF | High | Vericiguat, a soluble guanylate cyclase stimulator, did not significantly reduce NT-proBNP levels after 12 weeks but was well-tolerated. |

Table S2. Odds ratio and ranking of comparison for Overall Survival

|  | Placebo | Pracliciguat | Riociguat | Vericiguat |
| --- | --- | --- | --- | --- |
| Placebo | Placebo | 1.05 (0.02, 60.97) | 3.69 (0.53, 29.15) | 0.42 (0.12, 1.16) |
| Pracliciguat | 0.95 (0.02, 58.35) | Pracliciguat | 3.54 (0.04, 348.08) | 0.39 (0.01, 26.68) |
| Riociguat | 0.27 (0.03, 1.88) | 0.28 (0, 24.88) | Riociguat | 0.11 (0.01, 0.97) |
| Vericiguat | 2.41 (0.86, 8.51) | 2.59 (0.04, 180.88) | 9.07 (1.03, 101.89) | Vericiguat |

Figure S1. SUCRA Ranking for Overall Survival

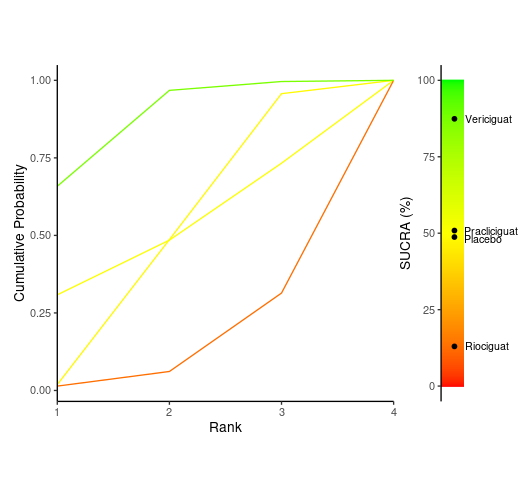

Table S3. Mean Difference and SD for LVEF change in the given studies

|  | Placebo | Pracliciguat | Riociguat | Vericiguat |
| --- | --- | --- | --- | --- |
| Placebo | Placebo | -0.19 (-7.34, 6.84) | 3 (-1.15, 7.13) | 1.59 (-2.03, 5.17) |
| Pracliciguat | 0.19 (-6.84, 7.34) | Pracliciguat | 3.18 (-5, 11.48) | 1.78 (-6.16, 9.73) |
| Riociguat | -3 (-7.13, 1.15) | -3.18 (-11.48, 5) | Riociguat | -1.4 (-6.88, 4.05) |
| Vericiguat | -1.59 (-5.17, 2.03) | -1.78 (-9.73, 6.16) | 1.4 (-4.05, 6.88) | Vericiguat |

Figure S2. SUCRA ranking for LVEF Change

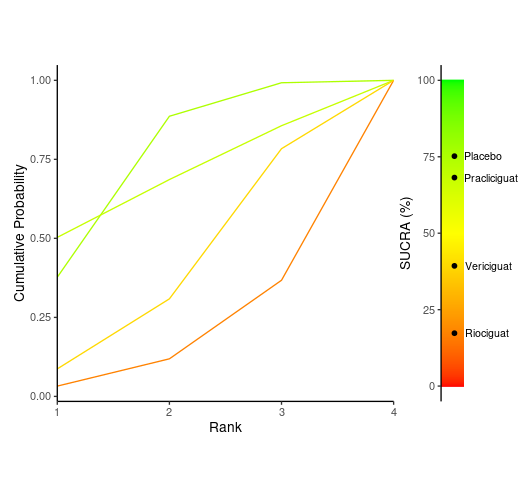

Table S4. Comparison and Treatment difference and ranking with Mean and SD for NT-proBNP

|  | Placebo | Pracliciguat | Riociguat | Vericiguat |
| --- | --- | --- | --- | --- |
| Placebo | Placebo | 5.28 (-12.58, 23.04) | -7.41 (-21.19, 4.41) | 0.64 (-9.54, 10.83) |
| Pracliciguat | -5.28 (-23.04, 12.58) | Pracliciguat | -12.68 (-35.54, 8.3) | -4.65 (-25.18, 15.87) |
| Riociguat | 7.41 (-4.41, 21.19) | 12.68 (-8.3, 35.54) | Riociguat | 8.04 (-7.51, 25.55) |
| Vericiguat | -0.64 (-10.83, 9.54) | 4.65 (-15.87, 25.18) | -8.04 (-25.55, 7.51) | Vericiguat |

Figure S3. SUCRA ranking for NT-proBNP

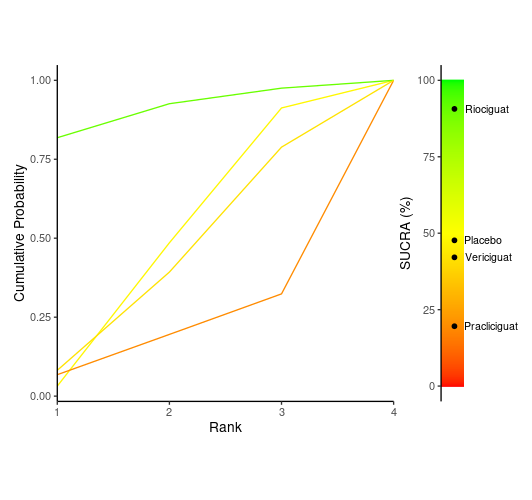

Table S5. Comparison and treatment difference in odds ratio for Rehospitalization

|  | Placebo | Riociguat | Vericiguat |
| --- | --- | --- | --- |
| Placebo | Placebo | 0.57 (0.12, 2.83) | 0.4 (0.11, 1.3) |
| Riociguat | 1.76 (0.35, 8.64) | Riociguat | 0.71 (0.09, 4.96) |
| Vericiguat | 2.48 (0.77, 9.33) | 1.4 (0.2, 11.73) | Vericiguat |

Figure S4. SUCRA Ranking for Rehospitalization

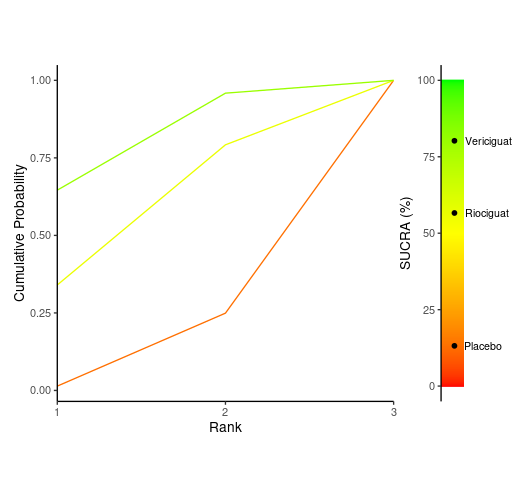

Figure S6. Risk of Bias

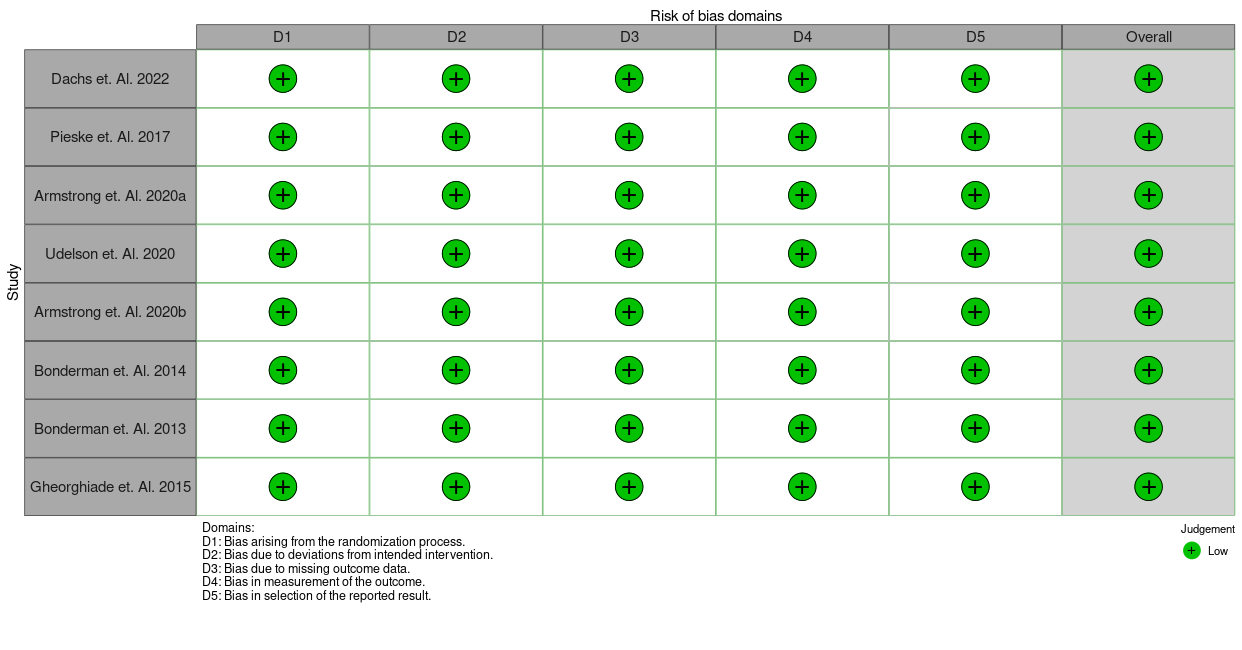
